# Unequal access to diagnosis of myalgic encephalomyelitis in England

**DOI:** 10.1101/2024.01.31.24302070

**Authors:** Gemma L. Samms, Chris P. Ponting

**Affiliations:** MRC Human Genetics Unit, The University of Edinburgh, Edinburgh, Scotland, EH4 2XU, UK

## Abstract

**Background:** People with Myalgic Encephalomyelitis (ME/CFS; sometimes referred to as chronic fatigue syndrome) experience very poor health-related quality of life and only rarely recover. ME/CFS has no curative treatment and no single diagnostic test. Public health and policy decisions relevant to ME/CFS require knowledge of its prevalence and barriers to diagnosis. However, people with ME/CFS report lengthy diagnostic delays and widespread misunderstanding of their symptoms. Published prevalence estimates vary greatly by country, gender, age and ethnicity.

**Methods:** Hospital Episode Statistics data is routinely collected by the NHS in England together with patient age, gender and ethnicity. This data, downloaded from the Feasibility Self-Service of NHS DigiTrials, was used to stratify individuals with the ICD-10 code that best reflects ME/CFS symptoms (G93.3; “Postviral fatigue syndrome”) according to their age, self-reported gender and ethnicity, General Practice and NHS England Integrated Care Board (ICB).

**Results:** In all, 100,055 people in England had been diagnosed with ME/CFS (ICD-10:G93.3) between April 1 1989 and October 7 2023, 0.16% of all registered patients. Of these, 79,445 were females and 20,590 males, a female-to-male ratio of 3.88:1. Female relative to male prevalence peaked at about 6-to-1 in individuals’ fourth and fifth decades of life. Prevalence varied widely across the 42 ICBs: 0.086%-0.82% for females and 0.024%-0.21% for males. White individuals were approximately 5-fold more likely to be diagnosed with ME/CFS than others; black, Asian or Chinese ethnicities are associated with particularly low rates of ME/CFS diagnoses. This ethnicity bias is stronger than for other common diseases. Among active English GP practices, 176 (3%) had no registered ME/CFS patients. Eight ICBs (19%) each contained fewer than 8 other-than-white individuals with a G93.3 code despite their registers containing a total of 293,770 other-than-white patients.

**Conclusion:** Those who are disproportionately undiagnosed with ME/CFS are other-than-white ethnic groups, older females (>60y), older males (>80y), and people living in areas of multiple deprivation. The lifetime prevalence of ME/CFS for English females and males may be as high as 0.92% and 0.25%, respectively, or approximately 390,000 UK individuals overall. This improved estimate of ME/CFS prevalence allows more accurate assessment of the socioeconomic and disease burden imposed by ME/CFS.

## Introduction

At the population scale, estimates of disease morbidity and prevalence inform public health and policy decisions; for individuals, accurate diagnosis informs symptom management and treatment. Myalgic Encephalomyelitis (ME; sometimes referred to as Chronic Fatigue Syndrome [1∼) is a chronic multi-system disorder resulting in very poor health-related quality of life and from which patients only rarely recover [2][3]. ME/CFS is relatively common [4]: prevalence estimates in the UK or the USA range from 0.2%-0.4% [5][6][7], depending on case definition. A meta-analysis of 56 data sets covering 1.1m individuals indicated an even higher prevalence, at 0.89% (95% CI: 0.60-1.33). However, this study’s criteria did not require post-exertional malaise (PEM), the worsening or new appearance of symptoms after previously tolerated physical or cognitive exertion, which nevertheless is a primary symptom of ME/CFS [8]. More recently, the USA National Health Interview Survey reported 1.3% of adults having a ME/CFS diagnosis and ongoing symptoms during 2021-2022 [9], and the All of Us study report 0.46% of their USA cohort with ME/CFS (ICD9CM-780.71; [10]).

ME/CFS diagnosis varies firstly by sex, with women diagnosed two- to five-times more often than men and also tending to report more severe symptoms [11][9]. Diagnosis also may vary by ethnicity, although studies disagree on which group is more frequently diagnosed: African Americans and Native Americans [12], or White or Black (non-Hispanic) individuals [9], or African American and Latinx youth [13][7].

In this study, we report Hospital Episode Statistics (HES) data for NHS Hospitals in England, specifically the ICD-10 code G93.3 (“Postviral fatigue syndrome”) which is the code that best reflects ME/CFS symptoms. Rather than being General Practice (GP) data, and rather than being UK-wide, the data is HES- and England-specific, and hence does not report UK-wide prevalence. Nevertheless, when stratified by age, self-reported ethnicity and sex the data reveals substantial population variation in ME/CFS diagnosis.

## Results

### Prevalence by age, deprivation and sex

On 7 October 2023, 100,055 people had an electronic health record code for ME/CFS across the 42 NHS England Integrated Care Boards (ICBs). These are among 62,782,175 people (0.16%) with a valid NHS number, who were registered with a GP and were not deceased. This information was supplied by the Feasibility Self-Service of NHS DigiTrials [14] who provide Hospital Episode Statistics that are routinely collected by the NHS in England and held by NHS England. In all, 79,445 (0.25% of 31,297,675) females and 20,590 (0.065% of 31,481,510) males were associated with the ICD10 G93.3 code, a female-to-male (F/M) ratio of 3.88:1; 20 people with ME/CFS did not specify their gender. F/M ratio exceeded 3.3 across all of the 42 ICBs (range 3.3-4.5) (**Figure 1A**).

**Figure 1.**
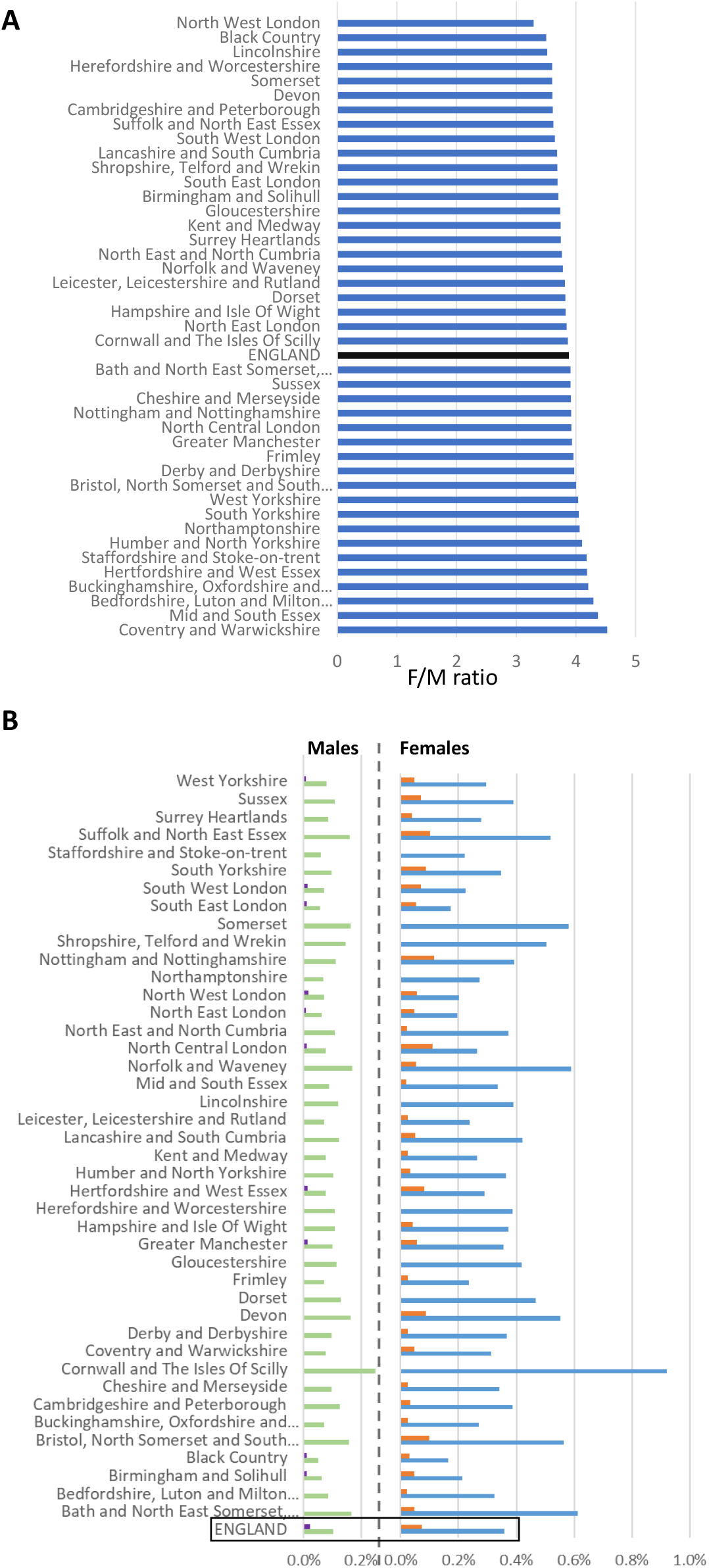
Diagnoses across 42 English Integrated Care Boards (ICB). (A) Ratios of female-to-male ME/CFS diagnoses (ICD-10:G93.3 codes) by ICB. (B) Point prevalence of males (left) or females (right) with ME/CFS stratified by ethnicity: either white or other-than-white. Blue: white females; Orange: Other-than-white Females; Green: white Males; Purple: Other-than-white Males.

Across the ICBS, female or male prevalence varied by an order of magnitude: 0.086%-0.82% and 0.024%-0.21%, respectively. Cornwall and the Isles of Scilly had the highest prevalence for each gender, and North West or North East London the lowest for females or males, respectively (**Figure 1B**).

Point prevalence of ME/CFS varied greatly across lifespan, peaking at about 50 years of age for females and over a decade later for males (**Figure 2A**). In individuals’ fourth and fifth decades the F/M ratio peaked at an exceptionally large value, approximately 6-to-1.

**Figure 2.**
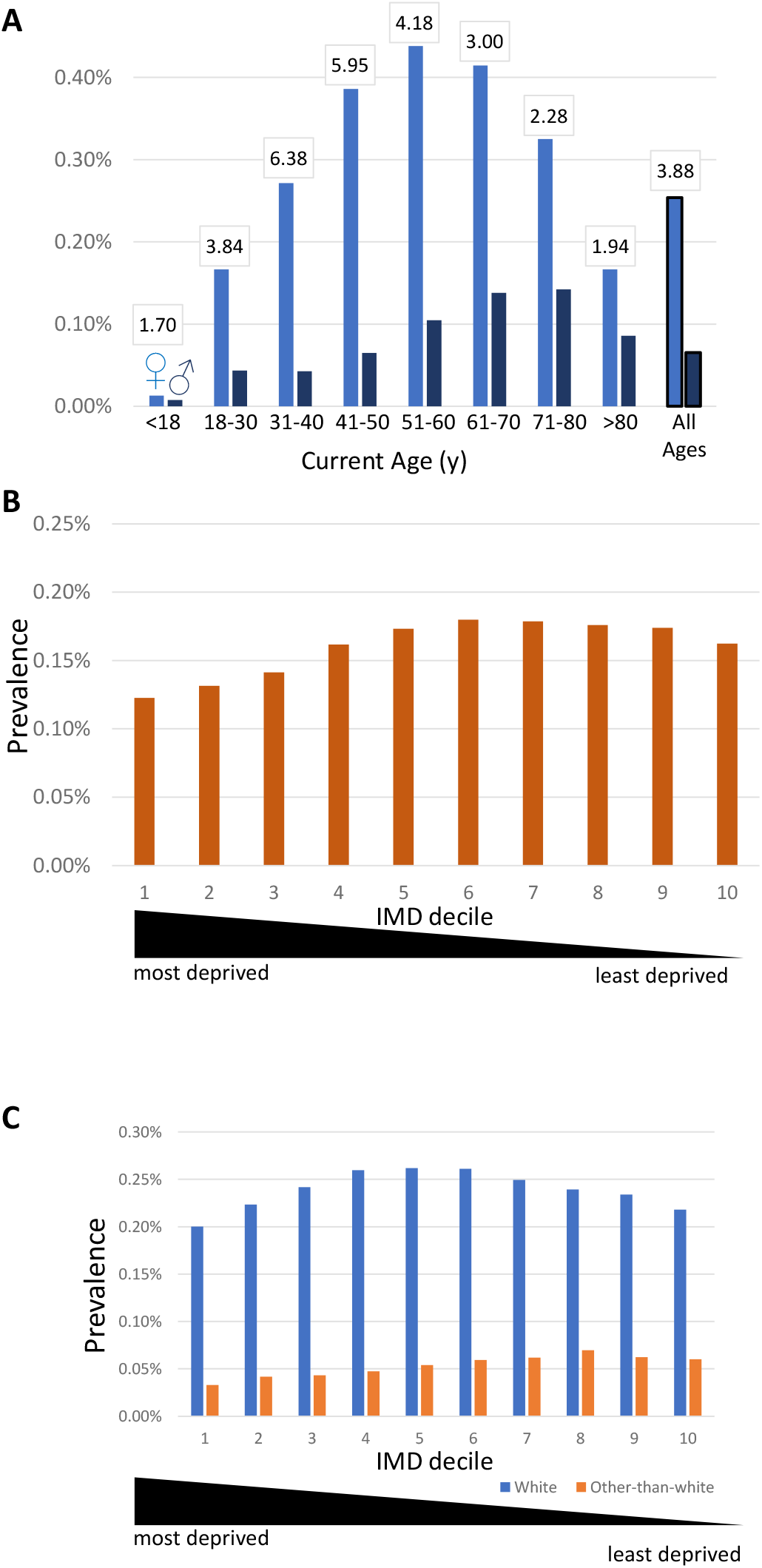
ME/CFS diagnoses by age or deprivation. (A) Ages of females or males with ME/CFS diagnoses in England (light blue: females; dark blue: males; black: overall ratio) as of 7^th^ October 2023. The female-to-male ME/CFS diagnosis ratios for every age decade are shown in boxes. (B) ME/CFS patients partitioned by Indices of multiple deprivation (IMD) in deciles. (C) As (B), but separated by self-reported white or other-than-white ethnicities.

ME/CFS prevalence varied 1.5-fold by deprivation assessed by the Index of Multiple Deprivation (IMD). The IMD is a measure of relative deprivation for small areas in England [15] combining seven domains: income; employment; education, skills and training; health and disability; crime; housing and services; and the living environment. Each small area has been ranked and divided into tenths (‘deciles’) from the most deprived (decile 1) to the least deprived (decile 10). ME/CFS prevalence was lowest for those living in areas among the three most deprived deciles (**Figure 2B**).

### Prevalence by ethnicity

Ethnic groups showed unexpectedly large variation in ME/CFS prevalence. Specifically, white prevalence was 4.9-fold higher than other-than-white prevalence (0.24% vs 0.049%, respectively). This finding was observed across all ICBs for both sexes (**Figure 1B**). Variation of ME/CFS prevalence by IMD deciles was similar between white and other-than-white ethnicities (**Figure 2C**).

ME/CFS prevalence of other-than-white British individuals was between one-tenth and one-third that for white British. Those with Chinese, Asian/Asian British or black/black British ethnicity were 11%, 11%-19%, or 10%-35%, respectively, less likely to be diagnosed with ME/CFS (**Figure 3A**). The low prevalence of ME/CFS for other-than-white ethnicity categories is more profound than for other diseases, for example fibromyalgia and clinical depression (**Figure 3A**).

**Figure 3.**
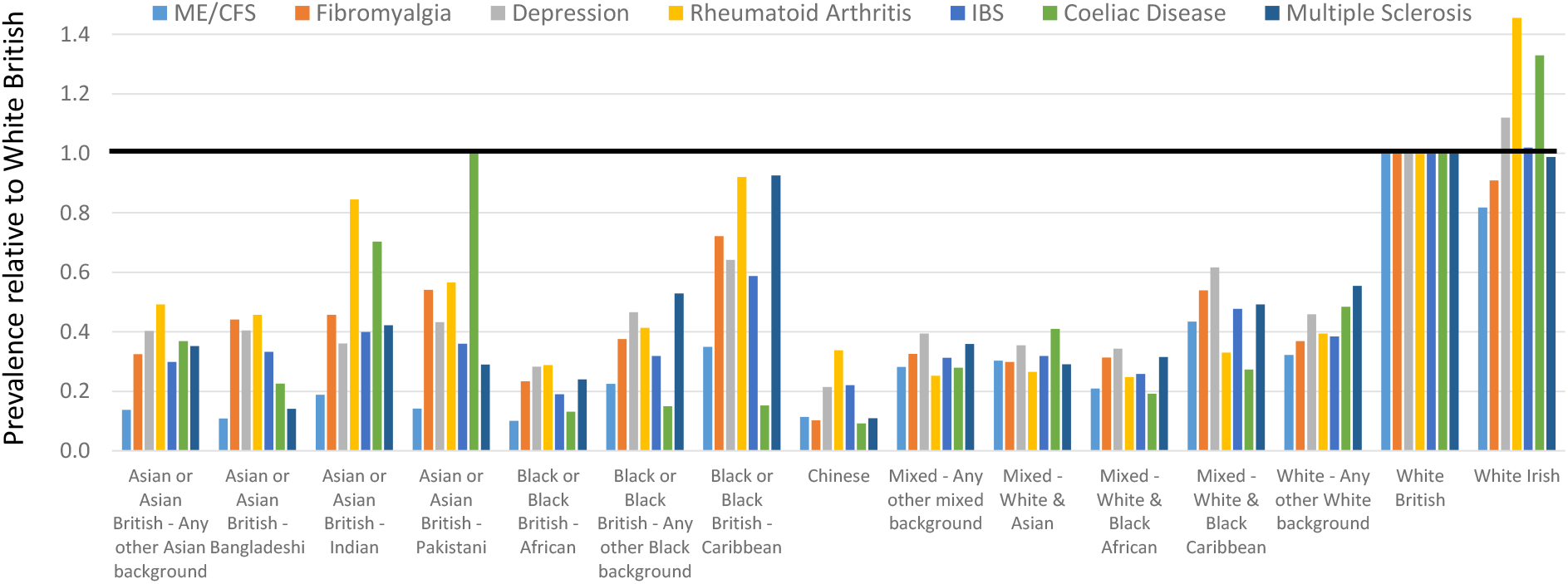
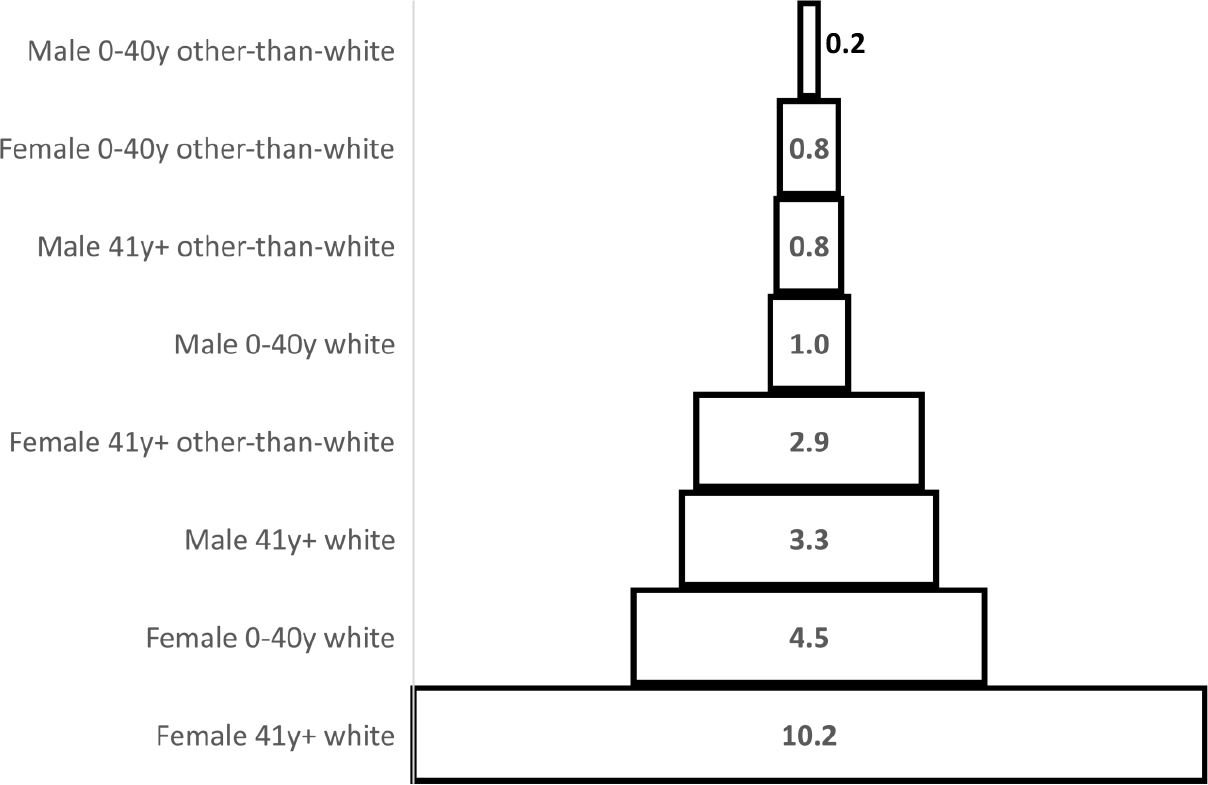
Relative prevalence of ME/CFS and other diseases stratified by ethnic group, age and gender. (A) Prevalence of ME/CFS (ICD-10:G93.3), Fibromyalgia (ICD-10: M79.7), Rheumatoid arthritis (M05.3, M05.8, M05.9, M06.0, M06.8, M06.9 or M08.0), clinical depression (F32 or F33 codes), Irritable Bowel Syndrome (K58.0 or K58.9), Coeliac Disease (K90.0), or Multiple Sclerosis (G35) for self-reported ethnicities relative to ‘white British’. Codes were downloaded from NHS DIgiTrials on 8^th^ October 2023. (B) ME/CFS prevalence for female or male groups, of younger or older age, and of white or other-than-white ethnicity, relative to young (≤40y old) white males (normalised to 1.0).

Eight ICBs each contained fewer than 8 other-than-white individuals with a G93.3 code (≤0.0027% for 293,770 other-than-white individuals). These 8 ICBs were Dorset; Gloucestershire; Herefordshire and Worcestershire; Lincolnshire; Northamptonshire; Shropshire, Telford and Wrekin; Somerset; and, Staffordshire and Stoke-on-Trent.

In summary, ME/CFS diagnosis in England is highest among older, white females, and lowest among younger other-than-white males; the variation is 50-fold between these three categories (**Figure 3B**).

### Prevalence per GP practice

Lastly, we considered 6,113 English GP practices with at least 2,500 registered patients which each, given the national prevalence of 0.16% (above), would be expected to have 4 or more ME/CFS patients registered. Two-thirds of these practices (4,061; 66%) had 8 or more registered patients with ME/CFS and 97% (5,937) reported at least one individual with ME/CFS. Of the remaining practices, 176 (3%) had no registered ME/CFS patients recorded in HES despite being operational and having a median of 4,765 registered patients (range 2,665-12,170), a total population of 917,570. These 176 practices are disproportionately located in the most deprived areas of England (**Figure 4**).

**Figure 4.**
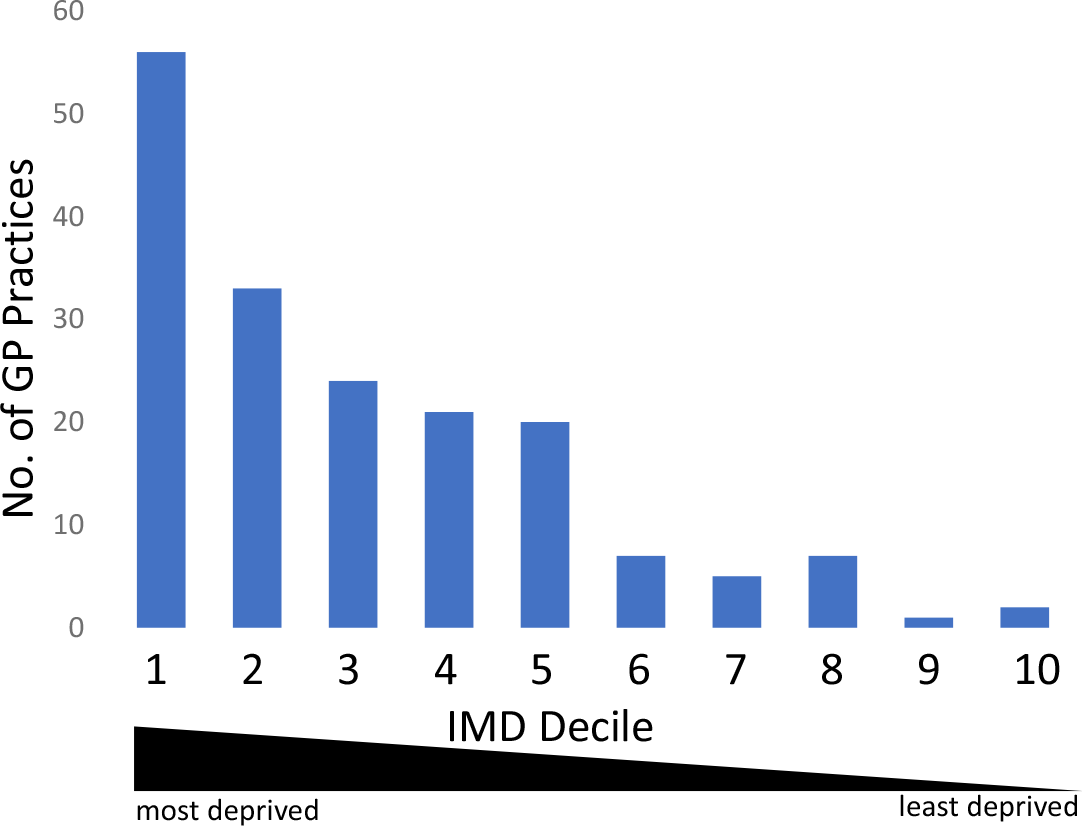
English GP practices. Numbers of active GP practices without registered ME/CFS patients recorded in HES stratified by indices of multiple deprivation.

## Discussion

Across England, ME/CFS diagnosis varies 50-fold, with considerable variation by gender, age, ethnicity, and location (**Figure 3B; Figure 1; Figure 2C**). Female-bias contributes about 4-fold, mid-way among previous values for single-country or international ME/CFS cohorts (1.5-6 fold [11][1][16][17][18][19][20]). This wide variation may result, in part, from cohorts sampling from different age ranges (**Figure 2A**). Young (<40y) or old (>40y) age contributes ∼3-fold to ME/CFS prevalence variation (**Figure 2A; Figure 3B**), reflecting an accumulation of ME/CFS diagnoses over adult life. Decreased ME/CFS female prevalence in later life (>60y; **Figure 1B**) cannot be due to recovery, as records are deleted only rarely, or increased mortality [21] but could reflect historically low levels of diagnoses. The 20y difference between the highest male (ages 71-80y) and highest female (ages 51-60y) prevalence is puzzling although it may reflect gender-specific historical variation in ME/CFS diagnosis. Older females (>60y) and males (>80y) are less likely to be coded with ME/CFS in their hospital records (**Figure 2A**). However, note that if they were diagnosed before 1st April 1989 and were not since re-coded in their hospital records, their diagnosis will be missing from the data we analysed.

White versus other-than-white ethnicity accounts for a further ∼5-fold variation (**Figure 3B**), with some ethnicities (e.g. black, Asian or Chinese ancestry) associated with particularly low rates of ME/CFS diagnoses (**Figure 3A**). This ethnicity bias is stronger than for other common diseases such as dementia, ischaemic heart disease, clinical depression, and fibromyalgia [22](**Figure 3A**). The bias is unlikely to reflect genetic differences [23], or to unequal access to primary care because other-than-white individuals are not less likely to visit GP services [24]. Rather it is likely due to fewer hospital visits by people of East Asian ancestry [24] and to wider social and economical health factors [25]. Barriers to diagnosing and managing ME/CFS in other-than-white groups have previously been recognised [26]. These will need to be overcome if there is to be equitable access to ME/CFS diagnosis and healthcare.

Age-at-ME/CFS diagnosis is highly variable, especially for paediatric ME/CFS cases: this study identified few <18y old ME/CFS cases in England (0.010% for both genders combined; **Figure 2A**), consistent with the UK overall [11]. In other countries, however, paediatric prevalence of ME/CFS is much higher [27], e.g., 0.75% in a USA community cohort [13]; 26% of Norwegian ME/CFS patients were younger than 20y old [17]. Paediatric ME/CFS referral services in England and the rest of the UK are limited [28].

A strength of this study is that it captures all ME/CFS diagnoses given the G93.3 ICD-10 code during admissions, outpatient appointments and historical Accident and Emergency attendances at English NHS hospitals up to October 2023. It is over two orders of magnitude larger in scale than a primary care study of 3 English areas [5] whose prevalence estimate (0.11-0.20%) is comparable to this study’s G93.3 prevalence (0.16%). A weakness is that those whose ME/CFS was diagnosed in primary care will only have been included if their G93.3 code was subsequently added to the HES data in secondary care.

With this study’s data, we can now estimate the number of people in the UK who would have a ME/CFS diagnosis (i.e. the G93.3 code) if there were minimal social and healthcare barriers to diagnosis. For this, we used the maximal ME/CFS prevalences for white females and males (0.92% and 0.25%, respectively), which are for NHS Cornwall and Isles of Scilly (**Figure 1B**). This ICB would be expected to have the highest fraction of diagnosed ME/CFS patients as it has the oldest population (average ∼45y [females] and ∼44y [males]), the highest F/M ratio (1.04) and, most strikingly, the lowest other-than-white population (2.0%). If all 67 million UK citizens [29] were to have a lifetime prevalence matching the point prevalence of NHS Cornwall and Isles of Scilly then 83,750 males and 308,200 females would be given a G93.3 ME/CFS diagnosis in their lifetime. These are lower bound estimates because no account is taken of those in NHS Cornwall and Isles of Scilly who have yet to receive a ME/CFS diagnosis. This total of 391,950 would be a 57% increase over a current prevalence estimate of 250,000 [30]. Even if half of these do not meet more stringent diagnostic criteria for ME/CFS (as previously [31][32]), then this reduction could be offset by those diagnosed in primary care, but not coded in the HES data analysed in this study. In summary, we suggest that the 2023 UK lifetime prevalence of ME/CFS diagnosed using stringent criteria is approximately 300,000 for females and 85,000 for males (i.e., 0.92% and 0.25%, respectively).

In summary, these results reveal deficiencies in ME/CFS diagnosis across different groups in England. To address these deficiencies, improved training of medical professionals should be available [33] and research into identifying accurate diagnostic tests should be prioritised. Even when diagnosed, there is no curative therapy for ME/CFS, only symptom management [33] Nevertheless, an individual’s ME/CFS diagnosis provides considerable value as it: (i) counters the delegitimization of problems often experienced by ME/CFS patients [34][35], (ii) improves patient/professional relationships, (iii) facilitates symptom management, (iv) enables application for disability benefits, (v) assists recruitment to future clinical trials of potentially effective treatments, and (vi) helps individuals to join supportive and informative patient communities.

## Methods

The NHS DigiTrials Feasibility Self-Service (England only [14]) returned all ME/CFS diagnoses since 1^st^ April 1989, stratified by ICB, sex, age and self-reported ethnicity (accessed on 7-10 October 2023). For each stratum, NHS DigiTrials data is rounded: counts equal to or exceeding 8 are rounded to the nearest increment of 5; counts equal to or less than 7 were not provided and so were set to zero. Data was for 62,782,175 individuals who are alive, have a valid NHS number and are registered with a GP in England. Data also includes those who previously met these criteria but are no longer residing in England [36]. Individuals are recorded once without duplicates. NHS DigiTrials returns data derived from Hospital Episode Statistics (HES) for admissions, outpatient appointments and historical Accident and Emergency attendances [36]. HES are linked via the patients’ registered GP practice, and these 6,119 GP practices are linked to 42 ICBs. 6 GP practices were not considered further because they had fewer than 2,500 registered patients. HES data is updated monthly, on the second day of each month. Personal demographic data is updated daily. Patients with multiple diagnoses of the same type/code are counted only once, specifically the latest occurrence. As in other studies (e.g., [17]) we consider G93.3 to be the ICD-10 code that best reflects ME/CFS symptoms.

We apply the UK Government’s recommendations for describing ethnicity [37]. NHS DigiTrials uses self-reported ethnicity as classified in the 2001 census. ‘White’ includes ‘white British’, ‘white Irish’ and ‘Any other white background’. ‘Other than white’ includes any other ethnic group; Asian or Asian British - Any other Asian background; Asian or Asian British - Bangladeshi; Asian or Asian British - Indian; Asian or Asian British - Pakistani; Black or Black British - African; Black or Black British - Any other Black background; Black or Black British - Caribbean; Chinese; Mixed - Any other mixed background; Mixed - White & Asian; Mixed - White & Black African; Mixed - White & Black Caribbean). These two categories exclude those whose ancestry is ambiguous (i.e. ‘not stated’ or ‘unknown’). Individuals whose ethnicity was ‘not stated’ or ‘unknown’ were included for non-ethnicity related analyses.

NHS DigiTrials provided numbers of ME/CFS patients coded with G93.3 in active GP practices in England. For each GP practice these were indicated as between 1 and 7, or if equal to or above 8, provided to the nearest increment of 5. Active practices were those without a closed date, with ≥2,500 registered patients, whose status is ‘active’ and ‘operational’ (similarly for its related ICB), and with a valid postcode.

For Figure 2B and 2C patients were partitioned into deciles of English indices of deprivation (IMD; 2019) by their address (1=most deprived, 10=least deprived). For Figure 4, English indices of deprivation for GP practices were acquired using their postcodes [15].

## Data Availability

With permission from the Feasibility Self-Service of NHS DigiTrials all data will be available following peer-reviewed publication.

## Acknowledgements

NHS data is copyright © 2022, NHS England, re-used with the permission of NHS England. All rights reserved. Access to this data was provided by NHS DigiTrials and the Feasibility Self-Service. Funding for access was provided by the National Institute for Health and Care Research (NIHR) and Medical Research Council (MRC), grant number MC_PC_20005. GLS was funded by a PhD studentship from ME Research UK. We would like to thank Profs Sarah Wild and Raj Bhopal (University of Edinburgh) and Simon McGrath for helpful discussions, and Dr Diana Garcia (University of Edinburgh) for assistance in acquiring the NHS DigiTrials data.

